# Detecting Cerebral Ischemia from Electroencephalography During Carotid Endarterectomy Using Machine Learning

**DOI:** 10.1101/2023.10.04.23295638

**Authors:** Amir I. Mina, Jessi U. Espino, Allison M. Bradley, Parthasarathy D. Thirumala, Kayhan Batmanghelich, Shyam Visweswaran

## Abstract

Intraoperative stroke is a major concern during high-risk surgical procedures such as carotid endarterectomy (CEA). Ischemia, a stroke precursor, can be detected using continuous electroencephalographic (cEEG) monitoring of electrical changes caused by changes in cerebral blood flow. However, monitoring by experts is currently resource-intensive and prone to error. We investigated if supervised machine learning (ML) could detect ischemia accurately using intraoperative cEEG. Using cEEG recordings from 802 patients, we trained six ML models, including naïve Bayes, logistic regression, support vector classifier, random forest (RF), light gradient-boosting machine (LGBM), and eXtreme Gradient Boosting with random forest (XGBoost RF), and tested them on a validation dataset of 30 patients. Each cEEG recording in the validation dataset was labeled independently by five expert neurophysiologists who regularly perform intraoperative neuromonitoring. We did not derive consensus labels but rather evaluated an ML model in a pairwise fashion using one expert as a reference at a time, due to the experts’ variability in label determination, which is typical for clinical tasks. The tree-based ML models, including RF, LGBM, and XGBoost RF, performed best, with AUROC values ranging from 0.92 to 0.93 and AUPRC values ranging from 0.79 to 0.83. Our findings suggest that ML models can serve as the foundation for a real-time intraoperative monitoring system that can assist the neurophysiologist in monitoring patients.

## Introduction

The risk of perioperative stroke, which occurs during or within 30 days of surgery, has increased over the past 10 years despite improvements in medical optimization and surgical technique^1^. The incidence of perioperative stroke is associated with high mortality and morbidity, leading to detrimental long-term effects on patient health and quality of life. Most perioperative strokes, which affect about 50,000 patients annually in the US, happen during surgery^2^. The incidence of stroke during surgery is as high as 6% for cardiovascular and neurological procedures and as low as 1% for general surgical procedures^2^. Cardiovascular and neurological procedures make up about one million of the 50 million surgical procedures done annually in the US. Reduction in the risk of perioperative stroke would significantly improve patient safety and ease the financial burden.

During surgery, continuous blood flow to the brain is essential for brain function, as it ensures the delivery of oxygen and nutrients. Because of the brain’s limited capacity to store oxygen and nutrients, even brief cerebral blood flow (CBF) disruptions can cause debilitating brain damage^3^. Initially, when CBF is reduced or stopped, it results in reversible brain damage from ischemia^4^. If CBF is not restored rapidly, the brain damage becomes irreversible due to the death of brain cells and results in a stroke. Therefore, early detection of ischemia can lead to timely preventive measures, lowering the risk of stroke and permanent brain damage.

Electroencephalography (EEG) can continuously and non-invasively measure brain function. The EEG reflects alterations in the metabolic and electrical activity of cortical neurons that accompany decreased CBF^5^. Animal studies show that when CBF falls below 15–18 ml/100 g/min, it rapidly affects electrical activity and leads to “electrical failure,” which can be reversed by increasing CBF. This corresponds to ischemia, resulting in reversible brain damage. A persistent decrease in CBF to 10-12 ml/100gm/min causes “membrane failure,” which ultimately leads to cell death. This corresponds to a stroke, resulting in irreversible brain damage. Depending on the severity of the reduction in CBF, the transition from ischemia to stroke may occur within minutes or take several hours.

Intraoperative neurophysiological monitoring (IONM) with EEG is routinely used in the operating room to detect ischemia during high-risk procedures such as carotid endarterectomies (CEAs)^6^. CEA is a surgical procedure that removes plaque inside the common or internal carotid artery. Carotid artery clamping is a necessary component of the surgical procedure. At the time of clamping, and shortly thereafter, the brain is most susceptible to ischemia. Beyond CEA, patients who undergo high-risk cardiovascular and neurological surgery are increasingly monitored with intraoperative EEG because of the risk of cerebral ischemia. However, human visual monitoring is susceptible to error and requires considerable skill and resources. This limits the human monitoring of EEG during surgery. Monitoring intraoperative ischemia can be aided by computer-based methods employing machine learning (ML) to detect ischemia rapidly and accurately on EEG. In this paper, we describe the training of supervised ML models on a large dataset of intraoperative EEGs performed for CEAs and report the results of the evaluation of these models on an independent dataset.

## Background

In this section, we provide brief descriptions of the EEG and quantitative parameters derived from it, EEG changes related to ischemia, and the application of ML to EEG.

### EEG and continuous EEG

The EEG measures and records the brain’s electrical activity in terms of voltage over time, and the recording is obtained by attaching electrodes to standard locations on the scalp. IONM typically utilizes eight electrodes attached to four locations on the left side and four on the right side of the head. Each electrode is identified by a letter and a number. The letter indicates the region of the brain beneath the electrode, such as F for the frontal lobe, P for the parietal lobe, T for the temporal lobe, and O for the occipital lobe. Even numbers represent the right side of the head, whereas odd numbers represent the left. Consequently, F3 is an electrode location above the brain’s left frontal lobe, while O2 is an electrode location above the right occipital lobe. Voltage is typically measured between two electrodes and displayed as a channel. Consequently, the F3 – P3 channel measures the voltage across the F3 and P3 electrodes (Figure 1). In typical IONM, eight distinct channels are displayed, each displaying the voltage difference between two electrodes comprising that channel (Figure 1). Technological advancements in the 1990s enabled the EEG to be digitized and recorded in electronic data storage, allowing continuous EEG (cEEG) recordings over hours.

**Figure 1.**
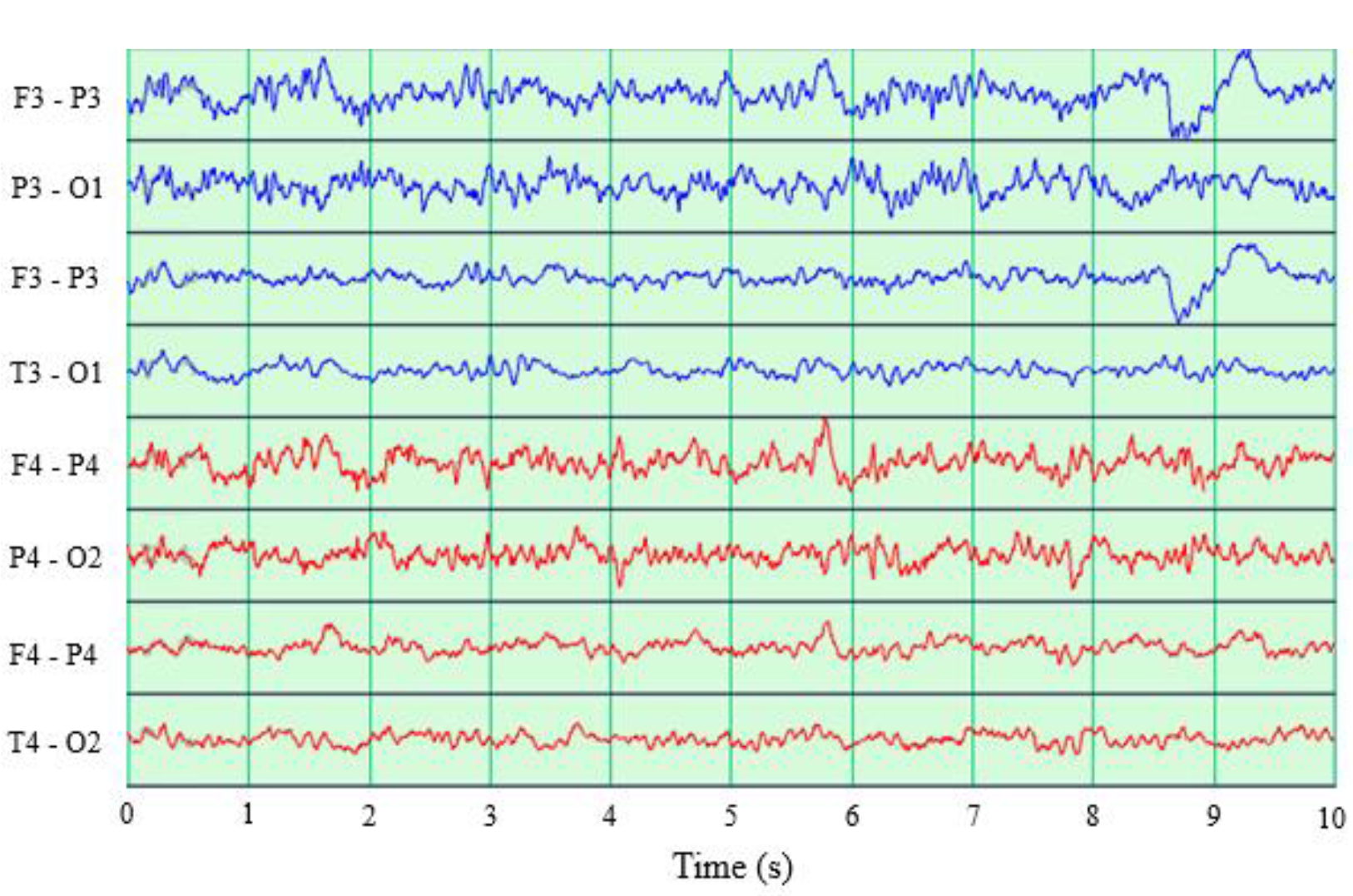
An eight channel EEG segment is shown. Channels F3-P3, P3-O1, F3-T3, and T3-O1 (blue) are for the left side of the head, while channels F4-P4, P4-O2, F4-T4, and T4-O2 (red) are for the right side.

In the operating room for the past few decades, cEEG has been used by anesthesiologists to monitor the depth of anesthesia and by neurophysiologists to monitor cerebral ischemia. Visual interpretation focuses on amplitude and morphology changes in EEG waveforms over time in specific frequency bands^4^; the clinically used frequency bands are delta (0 – 4 Hz), theta (4 – 8 Hz), alpha (8 – 12 Hz), beta (12 – 30 Hz), and gamma (30 – 100 Hz) in increasing order of frequency. Typically, as the frequency increases, the amplitude of the EEG signal decreases. Thus, the delta and theta bands have lower frequencies but larger amplitudes (and are referred to as slow activity), while the alpha, beta, and gamma bands have higher frequencies but smaller amplitudes (and are referred to as fast activity).

### Quantitative EEG

Performing visual analysis of the cEEG for extended periods of time in real-time is challenging. Furthermore, such analysis requires practice, and subtle changes in pattern can be missed with only visual inspection. Consequently, quantitative parameters for detecting ischemia have been developed. These parameters are referred to as quantitative EEG (qEEG) and are computed by mathematically processing digitally recorded EEG to identify patterns, quantify variations, and compare them to norms. Two types of qEEG parameters exist: those based on the frequency and those based on the amplitude of EEG signals. Frequency-based qEEGs are typically derived from the power spectrum obtained by applying fast Fourier transform (FFT) to the EEG signals. The power of a clinically used EEG band (e.g., the alpha band) is computed by aggregating the power over the specified frequency range (e.g., 8 – 12 Hz). Additionally, ratios of power in a higher frequency band to power in a lower frequency band have been found to be useful. Power ratios include the alpha-to-delta ratio (ADR), the beta-to-delta ratio (BDR), and the alpha-beta to delta-theta ratio (ABDTR)^7^. Another parameter is the spectral edge frequency (SEF) which is the frequency below which a specified proportion (often 90%) of the power in the EEG signal is contained^8^. Since ischemia and stroke typically affect one-half of the brain, parameters that measure the degree of asymmetry in brain activity have been developed, and one such qEEG is the pairwise-derived brain symmetry index (pd-BSI). One commonly used amplitude-based parameter is the amplitude-integrated EEG (aEEG), the amplitude of filtered and smoothed EEG signals^9^. Table 1 summarizes the qEEG parameters that are useful in ischemia.

**Table 1.** Selected qEEG parameters with descriptions that are used in ischemia detection.

| Type of qEEG parameter | qEEG parameter | Description |
| --- | --- | --- |
| Power | Delta power<br>Theta power<br>Alpha power<br>Beta power<br>Gamma power | The power or amount of activity in the specified frequency band in an EEG segment. |
| Power ratio | Alpha-to-delta ratio (ADR)<br>Beta-to-delta ratio (BDR)<br>Alpha-beta to delta-theta ratio (ABDTR) | The ratio of powers in two specified frequency bands in an EEG segment; often, the numerator is the power of a higher frequency band, and the denominator is the power of a lower frequency band. |
| Spectral edge frequency | Spectral edge frequency 90 (SEF90) | Spectral edge frequency is the highest frequency below which a specified amount of power is present in an EEG segment. SEF90 is the frequency below which 90% of the power is present. |
| Pairwise-derived brain symmetry index (pd-BSI) |  | The difference in power between matching electrode pairs on the left and right hemispheres, summed across frequencies. |
| Amplitude-integrated EEG (aEEG) |  | The range of the amplitude (difference between highest and lowest voltages) in an EEG segment. |

### EEG changes related to ischemi

Ischemic changes that occur during CEA have been documented in the literature. Visser et al. analyzed data from 92 CEA patients who, in addition to cEEG monitoring, underwent transcranial Doppler ultrasonography to measure blood flow velocity in intracranial arteries^10^. Following clamping, severe ischemia was associated with a reduction in alpha and beta band activity followed by an increase in delta and theta band activity, whereas mild ischemia was only associated with a reduction in alpha band activity. Some patients exhibited an increase in fast activity (alpha and beta) and a decrease in slow activity (delta and theta), which was attributed to arousal brought on by pain and hemodynamic stress caused by clamping. Kamitaki et al. analyzed cEEG recordings of 118 CEA patients and found that decreases in alpha, beta, and theta power of 52.1%, 41.6%, and 36.4%, respectively, were the strongest predictors of ischemia following clamping^11^. Pedapati et al. investigated the utility of 10 QEEG parameters in detecting cerebral ischemia during CEA using a database of 1,551 patients and found a statistically significant decrease in the trough values in all 10 qEEG parameters (alpha, beta, theta, delta, and gamma powers, ADR, BDR, ABDTR, SEF90, and aEEG) when there were visual changes in the EEG^12^.

### Machine learning in EEG

Automatic EEG analyses have extensively used ML for tasks such as epileptic seizure detection and prediction, sleep staging, and the diagnosis of Alzheimer’s disease, depression, and traumatic brain injury. While ML has been utilized for detecting stroke on the EEG, few studies have examined the detection of ischemia on the EEG, and none have examined the detection of ischemia in the operating room. In one study, ML was used to predict stroke by analyzing 30-second EEG segments from 80 patients, of whom half had suffered an ischemic stroke. The results of the study showed that tree-based ML methods were more effective than a multi-layer perceptron (MLP), with the area under the receiver operating curve (AUROC) values of 1.00 and 0.85 respectively. However, the results are likely over-optimistic due to overfitting, as the stratification was done on an interval level rather than at the patient level^13^. Another study utilized logistic regression on EEG features to predict stroke, achieving an AUROC value of 0.78, which improved to 0.8 when clinical features were incorporated^14^. This study found an AUROC of 0.88 when deep learning was applied to a combination of clinical and EEG features, but three-based methods were not compared. As far as we know, no studies have investigated ischemia detection using intraoperative EEG.

## Methods

In this section, we describe the datasets used for the study, details of preprocessing, the ML methods, the experimental design, and evaluation.

### Datasets

We used a large dataset of patients who underwent CEA while being monitored by cEEG in a large academic health system. All cEEG data were collected at 500 Hz and comprised four channels F3-P3, P3-O1, F3-T3, and T3-O1 on the left side and the four channels F4-P4, P4-O2, F4-T4, and T4-O2 on the right side. The training dataset included cEEG recordings from 802 patients who underwent CEA between 2009 and 2017. The recordings were reviewed by a single expert neurophysiologist, who labeled the time periods in each recording that indicated ischemia based on the monitoring neurophysiologist’s documented comments in the operating room. To expediently label such a large dataset, we employed a single expert who relied on the documented comments to make determinations of when ischemia occurred. These labels are referred to as intraoperative labels. Of the 802 patients, 51 were determined to have experienced ischemic changes during one or more time periods within 10 minutes of clamping on the cEEG. Data were collected retrospectively from various hospitals affiliated with the University of Pittsburgh Medical Center, where IONM employing cEEG was performed. Local Institutional Review Board approval was obtained for the data compilation, deidentification, and subsequent analyses.

The evaluation dataset comprised an independent set of 30 CEA patients, half of whom exhibited ischemic changes on cEEG. Five expert neurophysiologists independently reviewed the recordings in this dataset. The five reviewers included one board-certified neurologist with fellowship training in IONM and four neurophysiology technologists with certification in neurophysiologic intraoperative monitoring; all reviewers had a minimum of three years of experience with cEEG monitoring during vascular surgeries. The reviewers were provided with electronic files of cEEG recordings, which they viewed using the same software they used in the operating room. However, patient identifying information, such as demographics and monitoring neurophysiologist’s comments, was not provided. Reviewers were given the time at which the carotid clamp was placed and instructed to carefully examine a 30-minute period of the cEEG beginning at clamp time. They could freely scroll forward and backward through the cEEG and provide the precise time intervals at which cerebral ischemia-related changes occurred. These labels are referred to as refined labels. We concentrated on obtaining labels for only 30 minutes post-clamp, as this is when nearly all ischemic changes occur during CEA. This expedited labeling for each patient’s cEEG recording, which lasted an average of several hours.

### Preprocessing

A 10-minute segment of the cEEG recording beginning at clamp time was extracted for each patient in the training dataset. The EEG was filtered with standard filters, including a high-pass filter at 0.1 Hz, a low-pass filter at 70 Hz, and a notch filter at 60 Hz. Each 10-minute section was divided into 20-second non-overlapping intervals, and from each interval, 111 qEEG-based features were computed for ML. The frequency-based features were derived from the power spectrogram computed by applying FFT to a sliding interval to determine the power of each frequency. The spectrogram’s power values were converted to decibel units by taking 10 times the natural logarithm of the original power values. For each of the eight EEG channels, we computed five band powers (delta, theta, alpha, beta, and gamma), three power ratios (ADR, BDR, and ABDTR), SEF90, and aEEG (see Table 2). Except for the aEEG, all features were derived from the power spectrum. In addition to the single-channel features, we computed 10 averaged features based on four channels on the left side of the head, four on the right side of the head, and all eight channels respectively (see Table 2). The final feature was pd-BSI, which is computed from the power of all channels across all frequency bands.

**Table 2.** Summary of 111 ML features computed from each 20-second interval.

| Type of feature | Number of features |
| --- | --- |
| Features derived from a single channel | 10 features x 8 channels = 80 features |
| Features derived by averaging 4 channels from one side of the head | 10 features x 2 sides = 20 features |
| Features derived by averaging all 8 channels | 10 features |
| pdBSI | 1 feature |

We normalized all features at the patient level to a 120-second pre-clamp EEG baseline segment. To identify a 120-second pre-clamp baseline EEG with minimal noise and variance for each patient, we searched all possible 120-second intervals that ended just before clamp time. We then estimated the baseline distribution parameters from this 120-second interval. Using the baseline distribution, we computed each feature’s mean and standard deviation and then used these statistics to transform each feature value into a z-score.

For each 20-second interval, we derived a positive (i.e., ischemic) or negative (i.e., non-ischemic) label. We identified positive intervals by superimposing their time frames with the expert-provided periods of ischemia. An interval was labeled positive if more than half of its duration coincided with a period of ischemia. We preprocessed the evaluation dataset similarly to the training dataset to obtain 111 features for each 20-second interval. In contrast to the training dataset, which contained only one intraoperative label per interval, we created five additional labels per interval for the evaluation dataset. Thus, each interval in the evaluation dataset had one intraoperative label and five refined labels.

### Machine learning methods

We applied several supervised ML methods to derive learning classification models, including naïve Bayes (NB), elastic-net logistic regression (LR), support vector classifier (SVC) with radial basis function kernel, random forest (RF), light gradient-boosting machine (LGBM), and eXtreme Gradient Boosting with random forest (XGBoost RF). We focused on tree-based ML methods since they have been shown to outperform other methods, including deep learning for tabular data^15^. Common tree-based methods are RF and gradient boosting methods such as LGBM^16^ and XGBoost^17^. Few studies compared tree-based and deep learning models for EEG data. Nevertheless, it appears that tree-based models perform better in the context of EEG data, such as when modeling drowsiness^18^.

### Experimental protocols

The ML models were trained and evaluated at a 20-second interval level. We used 10-fold cross-validation to derive the models and optimize the hyperparameters. The training dataset was randomly partitioned into ten folds, such that all 20-second intervals derived from the same patient were assigned to the same fold to prevent overfitting. Further, the folds were constructed such that the proportion of intervals was approximately the same. For each ML method, ten base models, one from each fold, were trained and combined to form a soft voting ensemble classifier. When applied to an evaluation interval, the ensemble classifier produces a single probability by averaging the probabilities generated by the ten base models. The base models were calibrated using isotonic scaling. We implemented the preprocessing pipelines in Python 3.7.12 and the ML methods with scikit-learn 1.2.0.

### Evaluation methods

We report the performance of the models on the validation set using the five sets of expert-refined labels. The performance metrics included threshold-based sensitivity and specificity values and the threshold-free AUROC and area under the precision-recall curve (AUPRC) values. We used a threshold of 0.5 for computing sensitivity and specificity values. The AUROC is the most popular threshold-free metric for evaluating ML models^19^. Nonetheless, the AUPRC is increasingly utilized, particularly for imbalanced datasets such as ours. AUROC measures a model’s ability to discriminate between positive and negative samples across different probability thresholds and is insensitive to the distribution of labels. AUPRC, on the other hand, measures the trade-off between precision and recall across different probability thresholds and is sensitive to the distribution of labels, making it useful for imbalanced datasets^20^. Specifically, it has been demonstrated that AUPRC is a more informative metric than AUROC in rare disease settings with a small proportion of positive labels. The precision-recall (PR) curve has been shown to overcome the optimism of the receiver operating characteristic (ROC) curve in such situations and provide a more realistic evaluation of model performance^21^.

The interrater agreement on the evaluation dataset was calculated using pairwise crude percent agreement and Cohen’s kappa. We computed the agreement of each expert’s refined labels with those of the remaining experts (n=4). The mean and standard deviation were computed from the pairwise agreement comparison containing that reviewer in the pair. Similarly, for each model, the mean and standard deviation were calculated from the pairwise agreement of the model predictions with the refined labels of each expert (n=5). We also computed the agreement of the intraoperative labels with each set of refined labels and model predictions. In contrast to agreement, the pairs used to compute the statistics are order dependent, as one expert in a pair is designated as the reference with the “ground truth” and the other is assessed against the reference expert. For each expert, the mean and standard deviation of the evaluation metrics were computed using the remaining experts as references (n=4), and for a model, the mean and standard deviation were computed using the experts as references (n=5). From the probabilities obtained by the application of a model on the evaluation dataset, that model’s receiver operating characteristic (ROC) curve and precision-recall (PR) curve were calculated using the respective average metrics (i.e., mean sensitivity and mean specificity for the ROC curve and mean precision and mean sensitivity (or recall) for the PR curve) at all possible thresholds applied to the probabilities. Given the small sample sizes (n=4 and n=5), we calculated 95% confidence intervals for the metrics using a t-distribution, which accounts for the increased variability and wider tails more appropriately than the normal distribution. Instead of using a z-score from the normal distribution, we used a t-value corresponding to the 95% confidence level and appropriate degrees of freedom.

## Results

The training dataset was comprised of 23,880 intervals derived from 802 patients, and 3.6% of intervals derived from 51 patients were labeled as ischemic. Thirty patients were included in the validation dataset, 15 with ischemic changes on cEEG (mean age 71.1 years, 47% female, 33% left CEA) and 15 without ischemic changes (mean age 65.2 years, 47% female, 53% left CEA). The validation dataset consisted of 900 intervals of which 368 intervals were labeled as ischemic by at least one of the five experts. Figure 2 shows the variance in the number of experts who concur that a given interval is ischemic. The experts agreed unanimously that 532 intervals were non-ischemic and 116 were ischemic. The remaining 252 intervals, however, exhibited a variable degree of agreement, with anywhere from one to four experts classifying them as ischemic.

**Figure 2.**
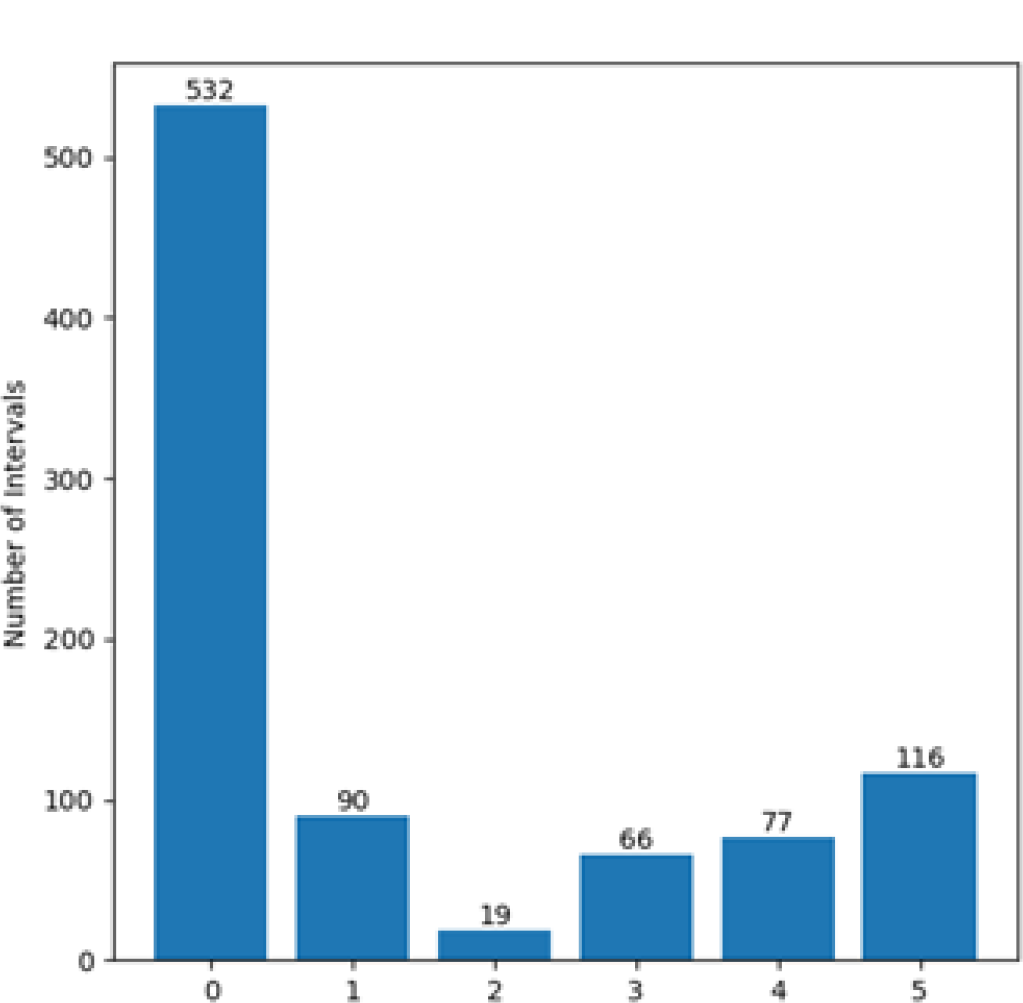
Histogram depicting the number of EEG intervals from the validation set labeled as ischemic by a varying number of experts. On the x-axis, a value of 0 indicates that all five experts agreed that the corresponding intervals were non-ischemic, a value of 1 indicates that at least one expert labeled the corresponding intervals as ischemic, and a value of 5 indicates that all five experts agreed that the corresponding intervals were ischemic.

### Agreement

The average agreements among experts and models are reported in Tables 2A and 2B. The experts’ mean Cohen’s kappa values ranged from 0.59 to 0.74, indicating moderate to substantial agreement. Except for NB and LR, mean Cohen’s kappa values for the models ranged from 0.61 to 0.66, which falls within the range of expert opinion. RF had the highest mean Cohen’s Kappa value among the models, at 0.66. In the case of NB, all predicted probabilities were smaller than 0.5, leading to all intervals being predicted as non-ischemic. The Cohen’s kappa coefficient adjusted for this by equating the expected and actual probabilities, resulting in a value of 0. Cohen’s kappa values assessing the agreement between intraoperative and refined labels ranged from 0.35 to 0.46, while those between the intraoperative labels and the model predictions (excluding NB and LR) ranged from 0.32 to 0.39.

**Table 2A.** Pairwise percent agreement and Cohen’s kappa for refined labels and percent agreement and Cohen’s kappa for intraoperative labels of the experts. Mean and SD refer to the mean and the standard deviation.

| Expert | Refined Labels |  | Intraoperative Labels |  |
| --- | --- | --- | --- | --- |
| | % Agreement (Mean ± SD) | Cohen's kappa (Mean ± SD) | % Agreement | Cohen's kappa |
| 1 | 0.84 ± 0.05 | 0.59 ± 0.09 | 0.76 | 0.35 |
| 2 | 0.87 ± 0.03 | 0.65 ± 0.04 | 0.76 | 0.37 |
| 3 | 0.90 ± 0.05 | 0.74 ± 0.13 | 0.75 | 0.39 |
| 4 | 0.85 ± 0.05 | 0.65 ± 0.13 | 0.76 | 0.46 |
| 5 | 0.89 ± 0.06 | 0.71 ± 0.16 | 0.74 | 0.36 |

**Table 2B.** Percent agreement and Cohen’s kappa for refined labels of the models and percent agreement and Cohen’s kappa for intraoperative labels of the models.

| Model | Refined Labels |  | Intraoperative Labels |  |
| --- | --- | --- | --- | --- |
| | % Agreement (Mean ± SD) | Cohen's kappa (Mean ± SD) | % Agreement | Cohen's kappa |
| NB | 0.73 ± 0.07 | 0.00 ± 0.00 | 0.71 | 0.00 |
| LR | 0.84 ± 0.04 | 0.54 ± 0.05 | 0.72 | 0.24 |
| SVC | 0.86 ± 0.02 | 0.63 ± 0.04 | 0.76 | 0.39 |
| RF | 0.86 ± 0.03 | 0.66 ± 0.08 | 0.72 | 0.36 |
| LGBM | 0.86 ± 0.03 | 0.61 ± 0.04 | 0.75 | 0.32 |
| XGBoost RF | 0.86 ± 0.02 | 0.65 ± 0.04 | 0.72 | 0.33 |

### Predictive performance

Figure 3 shows the ROC curves and PR curves for the models and a single operating point for each of the experts since they did not provide probabilities. The sensitivity and specificity values for experts and models and the AUROC and AUPRC values for the models are shown in Tables 3A and 3B. The experts’ mean sensitivity and specificity ranged from 58% to 93% and from 83% to 96%, respectively. Except for NB and LR, the models’ mean sensitivity and specificity ranged from 63% to 89% and from 85% to 96%, respectively. Again, with NB, all intervals were predicted as non-ischemic, resulting in a sensitivity of 0% and a specificity of 100%.

**Table 3A.** Mean sensitivity and specificity values with 95% confidence intervals for the five experts.

| Expert | Sensitivity | Specificity |
| --- | --- | --- |
| 1 | 0.58 (0.36, 0.80) | 0.96 (0.93, 0.99) |
| 2 | 0.66 (0.45, 0.86) | 0.96 (0.91, 1.00) |
| 3 | 0.90 (0.71, 1.00) | 0.91 (0.77, 1.00) |
| 4 | 0.93 (0.87, 0.99) | 0.83 (0.71, 0.94) |
| 5 | 0.81 (0.66, 0.96) | 0.92 (0.76, 1.00) |

**Table 3B.**
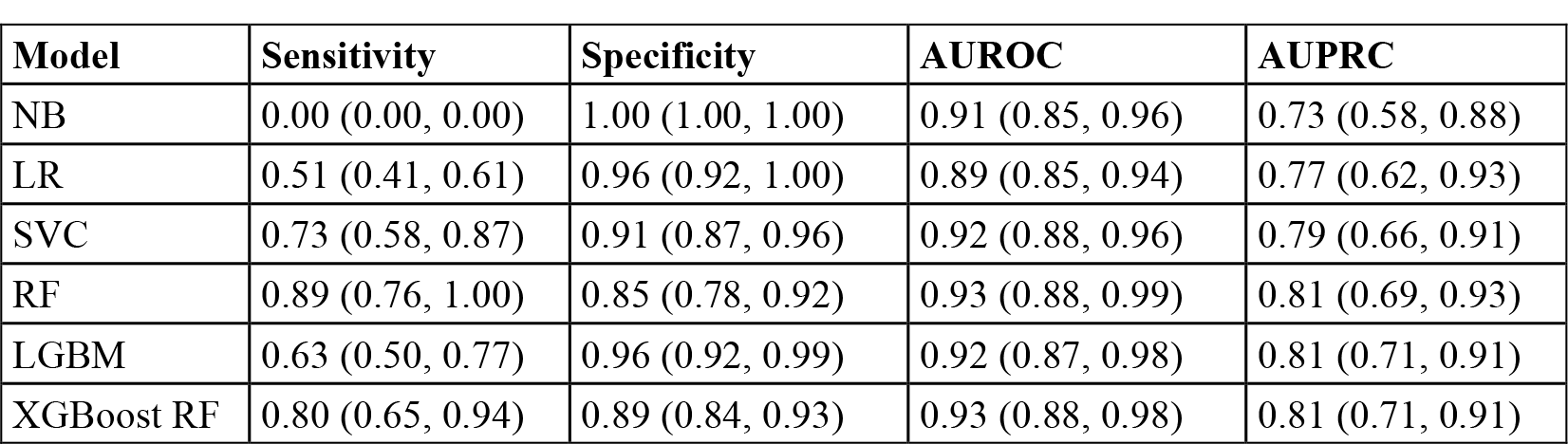
Mean sensitivity, specificity, AUROC, and AUPRC values with 95% confidence values for the six models.

| Model | Sensitivity | Specificity | AUROC | AUPRC |
| --- | --- | --- | --- | --- |
| NB | 0.00 (0.00, 0.00) | 1.00 (1.00, 1.00) | 0.91 (0.85, 0.96) | 0.73 (0.58, 0.88) |
| LR | 0.51 (0.41, 0.61) | 0.96 (0.92, 1.00) | 0.89 (0.85, 0.94) | 0.77 (0.62, 0.93) |
| SVC | 0.73 (0.58, 0.87) | 0.91 (0.87, 0.96) | 0.92 (0.88, 0.96) | 0.79 (0.66, 0.91) |
| RF | 0.89 (0.76, 1.00) | 0.85 (0.78, 0.92) | 0.93 (0.88, 0.99) | 0.81 (0.69, 0.93) |
| LGBM | 0.63 (0.50, 0.77) | 0.96 (0.92, 0.99) | 0.92 (0.87, 0.98) | 0.81 (0.71, 0.91) |
| XGBoost RF | 0.80 (0.65, 0.94) | 0.89 (0.84, 0.93) | 0.93 (0.88, 0.98) | 0.81 (0.71, 0.91) |

**Figure 3.**
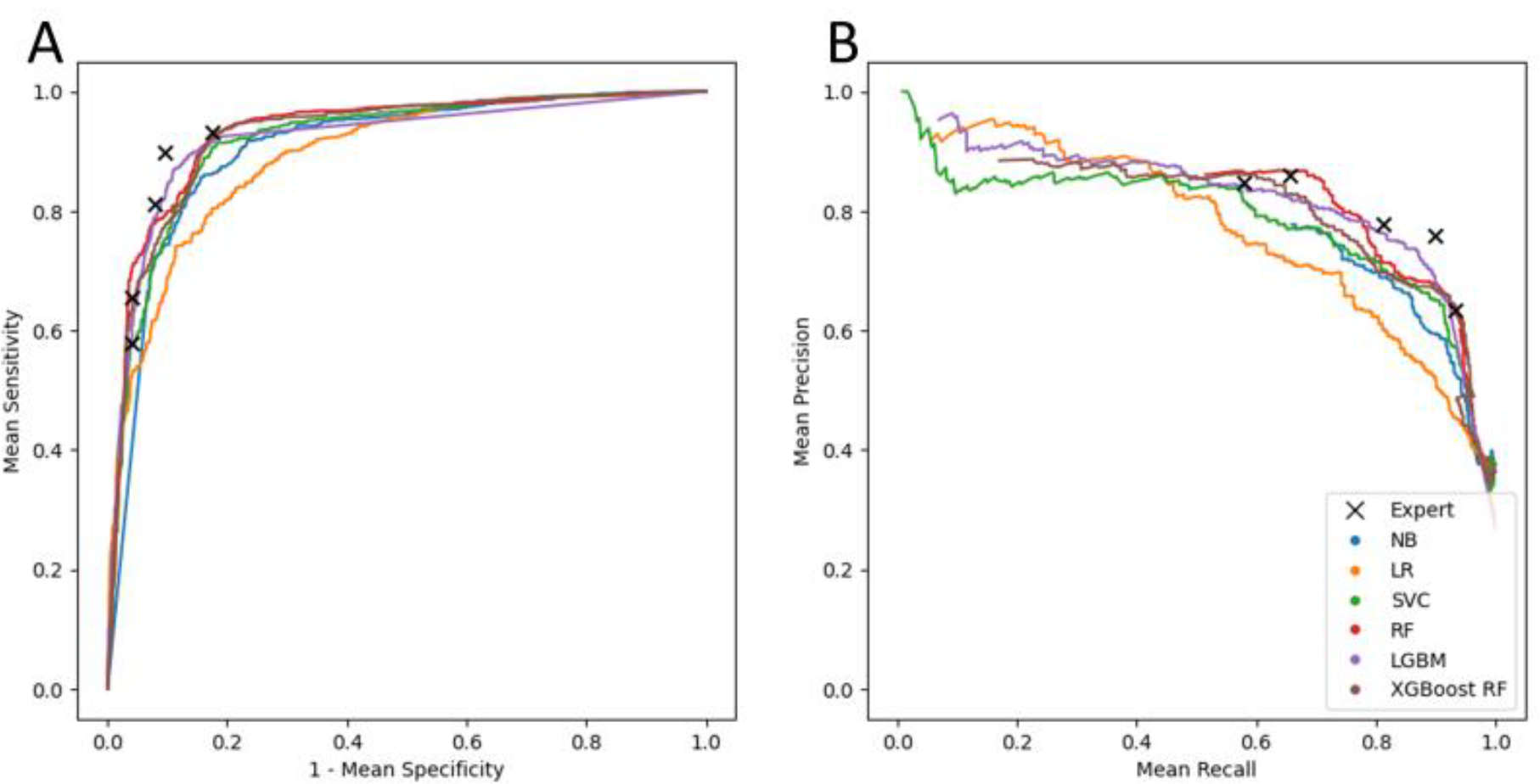
(A) ROC curves for the six models, as well as the mean sensitivity and specificity values for the five experts (multiplication symbols). (B) PR curves depicting the mean precision and recall values for the six models, as well as mean precision and recall values for the five experts (multiplication symbols).

## Discussion

Using a large training dataset of 802 cEEG recordings from CEA procedures, we developed multiple ML models for predicting intraoperative cerebral ischemia. The tree-based models including RF, LGBM, and XGBoost RF performed well on an independent validation dataset with AUROC values between 0.92 and 0.93 and AUPRC values between 0.79 and 0.83. The size of our dataset obtained from a single surgical procedure, namely CEA, which provides a clearly defined timeframe during which cerebral ischemia can occur, is a major strength of our study. Previous research has not looked at ML models for intraoperative cerebral ischemia, though a previous study examined the univariate correlations of qEEG features in CEA for detecting ischemia, but with a smaller sample of 118 patients^11^.

Another strength of our study was the use of an independent validation dataset in which each cEEG recording was carefully reviewed by five expert neurophysiologists. This allowed us to characterize expert variability in identifying time periods of EEG changes indicative of ischemia. Part of the variation is due to minor differences in the start and end times of the expert labels, even though most of them agreed that an ischemic change occurred during roughly the same time period. Furthermore, our analysis revealed that an expert review uncovered EEG changes that were not recorded intraoperatively, either because they were missed due to time constraints or because they were observed but not documented. We chose pairwise comparisons for evaluation over creating consensus gold standard labels due to the variability of the refined labels at the interval level. A similar approach has been used to evaluate other EEG-related tasks, such as seizure detection^22^.

In terms of sensitivity and specificity, the predictive performance of the tree-based models was comparable to the performance achieved by the experts when not under time constraints. Given the suboptimal performance of the experts during intraoperative procedures compared to their performance when not under time constraints, this supports the clinical use of ML-based models to alert the monitoring neurophysiologist.

### Limitations

Our study has several limitations. First, the cEEG recordings that we employed contained residual artifacts. Although we used band-pass and notch filtering to identify and remove some artifacts, others remained because no automated methods exist to identify them. Second, as described in the Methods section, for expediency, we used intraoperative documentation to derive labels for the training dataset, which we now know are suboptimal. Third, for modeling purposes, we ignored the time-series nature of the cEEG and treated each 20-second interval as statistically independent from the intervals that came before and after it. By incorporating temporal correlations, time series analysis and forecasting techniques such as autoregressive integrated moving average (ARIMA) may perform better. In addition, we did not evaluate deep learning ML models capable of handling time series data, such as recurrent neural networks and long short-term memory networks. Fourth, we employed standard atemporal evaluation metrics such as AUROC and AUPRC, whereas time-series aware metrics may provide a more precise evaluation of the model’s performance^23,24^.

### Future work

We intend to address many of the limitations described in the previous paragraph in future work. We are developing automated methods to identify EEG signal artifacts such as electrical artifacts that are common in the operating room and burst suppression artifacts that are common when general anesthesia is administered. We have begun obtaining refined labels for the training dataset, in which each cEEG recording is carefully reviewed by a single expert with no time constraints. Separately, we are investigating time-aware deep learning methods and adapting existing time-series aware metrics for model evaluation^25^.

## Conclusions

We derived several ML models for detecting cerebral ischemia from intraoperative cEEG and evaluated them on a validation set with labels refined by several experts. Though they were trained on cEEG recordings with artifacts and suboptimal labels, tree-based models such as RF, LGBM, and XGBoost RF performed well. Our results support that ML can be sufficiently accurate in monitoring cEEG in real time for intraoperative cerebral ischemia. Such an automated system can reduce intraoperative ischemia-related complications, improve patient safety, and reduce healthcare costs.

## Data Availability

All data produced in the present study are available upon reasonable request to the authors, consistent with IRB guidelines and data protection regulations.

## Acknowledgments

Research reported in this publication was supported by the National Institutes of Health under award number T32 GM008208 from the National Institute of General Medical Sciences, T15 LM007059 from the National Library of Medicine, and UL1 TR001857 from the National Center for Advancing Translational Sciences.

